# Immunophenotyping reveals longitudinal changes in circulating immune cells during radium-223 therapy in patients with metastatic castration-resistant prostate cancer

**DOI:** 10.1101/2020.11.11.20229831

**Authors:** J.H.A. Creemers, M.J. van der Doelen, S. van Wilpe, R. Hermsen, T. Duiveman-de Boer, D.M. Somford, M.J.R. Janssen, J.P.M. Sedelaar, N. Mehra, J. Textor, H. Westdorp

## Abstract

**Purpose:** Radium-223 improves overall survival (OS) in men with bone metastatic castration-resistant prostate cancer (mCRPC). While the exact mechanism behind this survival benefit remains unclear, radium-induced immunological mechanisms might contribute to the OS advantage. We performed a comprehensive evaluation of the immunological changes in mCRPC patients by phenotyping the peripheral blood mononuclear cells (PBMCs) during radium-223 therapy.

**Experimental Design:** In this prospective, single-arm, exploratory study, PBMCs of 30 mCRPC patients were collected before, during, and after treatment with radium-223. Lymphocyte and monocyte counts were analyzed to get insight into general immune cell trends. Next, we analyzed changes in T cell subsets, myeloid-derived suppressor cells (MDSCs), and immune checkpoint expression using linear regression models. Per subset, the 6-month change (% of baseline) was determined. Bootstrapped 95% confidence intervals were used to measure the degree of uncertainty of our findings.

**Results:** We observed a substantial decrease in absolute lymphocyte counts (−0.12 * 10^9 cells/L per injection, 95% CI: -0.143 - -0.102). Simultaneously, an increase was observed in the proportion of T cells that expressed costimulatory (ICOS) or inhibitory (TIM-3, PD-L1, and PD-1) checkpoint molecules. Moreover, the fraction of two immunosuppressive subsets – the regulatory T cells and the monocytic MDSCs – increased throughout treatment. These findings were not more pronounced in patients with an ALP response during therapy.

**Conclusion:** Immune cell subsets in patients with mCRPC changed during radium-223 therapy, which warrants further research into the possible immunological consequences of these changes.

## INTRODUCTION

Radium-223 dichloride (radium-223) was registered in 2013 to treat patients with symptomatic bone metastatic castration-resistant prostate cancer (mCRPC) based on the results of the phase III, randomized controlled ALSYMPCA trial. In this trial, radium-223 improved the overall survival (OS) and prolonged the time to a first symptomatic skeletal event [1].

Radium-223 is an alpha-emitting radionuclide that selectively binds to areas with increased bone turnover, such as bone metastases. Alpha particles are highly ionizing agents with low penetration power (≤100 µm) [2]. Their radiation induces double-stranded DNA breaks in adjacent tumor cells, osteoblasts, and osteoclasts, resulting in tumor cell death and inhibition of pathological bone formation [3,4]. In models of prostate cancer metastases, radium-223 was found to be deposited at the bone surface next to the tumor, not within the tumor itself [5]. It is mechanistically largely unknown how bone metastases respond to radium-223 and, therefore, it is unclear how radium-223 prolongs OS.

Immunological mechanisms may contribute to the OS benefit of radium-223. Preclinical studies have demonstrated that ionizing radiation triggers an immune response via the release of danger-associated molecular patterns (DAMPs), such as calreticulin. These DAMPs stimulate antigen-presenting cells to activate cytotoxic T lymphocytes, thereby inducing immunogenic tumor cell death [6]. A recent *in vitro* study indicates that radium-223, like radiation therapy, can also induce immunogenic modulation. Radium-223 enhanced T cell-mediated lysis of tumor cells through upregulation of major histocompatibility complex class I molecules and increased cell surface expression of calreticulin on tumor cells [7]. Nevertheless, data on the immunological effects of radium-223 in mCRPC patients is limited to one study that evaluated changes in circulating CD8^+^ T cells during the first 3-4 weeks of radium-223 therapy. This study in fifteen mCRPC patients observed a decrease in PD-1^+^ effector memory CD8^+^ T cells during treatment [8]. A better understanding of the immunological effects of radium-223 might provide a rationale for new treatment strategies, combining radium-223 with immune-based therapies, thereby possibly improving the clinical benefit of radium-223 therapy in mCRPC.

Here we investigated the composition and abundance of circulating peripheral blood mononuclear cells (PBMCs) of mCRPC patients collected before, during, and after treatment with radium-223. We postulated that changes in PBMCs could be seen during radium-223, as this has also been described in patients after external beam radiation [9]. A comprehensive, exploratory analysis was performed to investigate the longitudinal changes in T cell subsets, myeloid-derived suppressor cell (MDSC) subsets, and immune checkpoint expression during radium-223 therapy.

## METHODS

### Patient recruitment

This prospective, single-arm, exploratory study included mCRPC patients treated with radium-223 at the Radboudumc and Canisius-Wilhelmina hospital, Nijmegen, The Netherlands between May 2016 and January 2018. Patients were eligible if they had histologically proven adenocarcinoma of the prostate, mCRPC with symptomatic bone metastases, and no (history of) visceral metastases. Patients with a history of a secondary malignancy or auto-immune disease were excluded. Prior radionuclide treatment and concomitant other anticancer treatments were not allowed except for luteinizing hormone-releasing hormone agonists or antagonists. The use of low-dose corticosteroids (maximally 10 mg prednisone daily) was permitted.

Written informed consent was obtained from all patients before enrolment. The study was conducted in accordance with the principles of Good Clinical Practice, the Declaration of Helsinki, and other applicable local regulations and was approved by the institutional review boards of both participating centers.

### Radium-223 therapy and study-specific procedures

Patients were treated with radium-223 according to daily practice, with radium-223 being administered intravenously every four weeks at a dose of 55 kBq/kg, with a maximum of six injections. Laboratory evaluations, including lymphocyte and monocyte counts, were carried out within four weeks before radium-223 initiation, one week before each subsequent injection, and four weeks after the end of treatment. Within three months before the start of radium-223 therapy, bone scintigraphy, and CT or ^68^Ga-PSMA-11 PET/CT of the thorax, abdomen, and pelvis were performed. At baseline and four weeks after the second, fourth, and sixth radium-223 injection, 30ml of blood was collected in heparin blood collection tubes for immunophenotyping.

### Blood processing and storage

PBMCs were isolated using Ficoll-Paque gradient centrifugation. After adding Ficoll (Lymphoprep, Axis-Shield, Dundee, UK), samples were centrifuged at 2100 x g for 20 minutes at room temperature. The PBMC layer was transferred to a new tube and washed with phosphate-buffered saline (PBS). Viable cells were counted manually with a Bürker-Türk counting chamber. Cells were resuspended in culture medium (X-VIVO + 2% human serum) and mixed 1:1 with freezing medium (20% DMSO, 80% human serum albumin) at a concentration of 2-20 x10^6^ viable cells per ml. Cells were aliquoted into cryogenic vials and stored in liquid nitrogen.

### Flow cytometry

PBMCs were thawed rapidly in a 37 °C water bath and diluted in RPMI 1640 media. Cell number and viability were determined with a hemocytometer using trypan blue. Immunophenotyping of PBMCs was performed using four antibody panels. We used a T cell panel consisting of antibodies against CD3, CD4, CD8, CD25, CD45RO, CTLA-4 (CD152), CCR7 (CD197), and FoxP3 (Supplementary Table S1). In addition, we developed two checkpoint panels. Checkpoint panel 1 included antibodies targeting CD3, CD4, CD8, LAG-3 (CD223), PD-L1 (CD274), ICOS (CD278), PD-1 (CD279), and TIM-3 (CD366). Checkpoint panel 2 consisted of a CD3, CD4, CD8, OX-40 (CD134), CTLA-4, PD-L1, PD-1, and TIM-1 (CD365) antibody mix. Lastly, the MDSC panel consisted of antibodies against lineage markers (CD3, CD19, CD20, CD56), CD11b, CD14, CD15, CD33, PD-L2 (CD273), PD-L1, and HLA-DR. Antibody specifications are tabulated in Supplementary Table S1. In all panels, fixable viability dye efluor 780 (eBioscience, San Diego, CA, USA) was used to exclude dead cells.

Cells were incubated with fixable viability dye diluted in PBS at 4 °C for 30 minutes. Subsequently, cells were incubated with an antibody mix, consisting of the cell surface markers diluted in brilliant staining buffer (BD Biosciences, San Jose, CA, USA). The cells were incubated at 4 °C in the dark for 30 minutes. For intracellular staining, cells stained with the T cell panel were fixed with Fix/Perm (eBioscience) and incubated for 2 hours at 4 °C. After washing, the cells were resuspended in permeabilization buffer containing antibodies against the intracellular markers anti-FoxP3 and anti-CTLA-4 and incubated for 30 minutes at 4 °C.

Staining intensity was measured with the FACSLyric (BD Biosciences). Instrument settings were verified and adjusted before each acquisition using single stainings. Data were analyzed with FlowJo Software (Tree Star Inc., Ashland, OR, USA). Positive and negative cell populations for each marker were determined using fluorescence minus one (FMO) controls. The gating strategy is shown in Supplementary Figure S1. Regarding checkpoint expression, both the percentage of positive cells and the median fluorescence intensity (MFI) were studied. To minimize the effects of intra-batch differences, we used the ΔMFI, which is defined as the MFI of the test sample minus the MFI of the negative FMO control. Checkpoint molecules expressed on <1 % of the cell of interest were excluded from further analyses.

### Clinical response evaluation

Clinical response was defined as a decline in alkaline phosphatase (ALP) levels of ≥30% from baseline during therapy, according to the ALSYMPCA study criteria [1]. In addition, PSA responses, radiological responses, and OS were analyzed. PSA response was defined as a decline of ≥30% during treatment according to the ALSYMPCA study criteria. Within three months after completion or discontinuation of therapy, patients underwent radiological evaluation by bone scintigraphy and CT of the thorax, abdomen, and pelvis. Radiological evaluation of soft tissues was performed according to Response Evaluation Criteria In Solid Tumors version 1.1 [10], and bone scans were evaluated according to Prostate Cancer Working Group 3 criteria [11]. OS was defined as the time between the first radium-223 injection and either death from any cause or the last follow-up. All patients were followed until death or August 1, 2020.

### Data analysis

Exploratory data analyses were performed using R version 3.6.2. The following packages were used for analyses and visualization: the Tidyverse collection of packages [12], scales [13], cowplot [14], and ggbeeswarm [15].

Lymphocyte and monocyte counts were normalized to adjust for interindividual variation. Cell counts were normalized per patient as follows: the patients’ mean count per cell type was subtracted from the measurement, and, subsequently, the overall mean of the count of that particular cell type was added. Linear regression was used to determine the average linear change of the cell counts across radium-223 injections (Figure 1).

**Figure 1:**
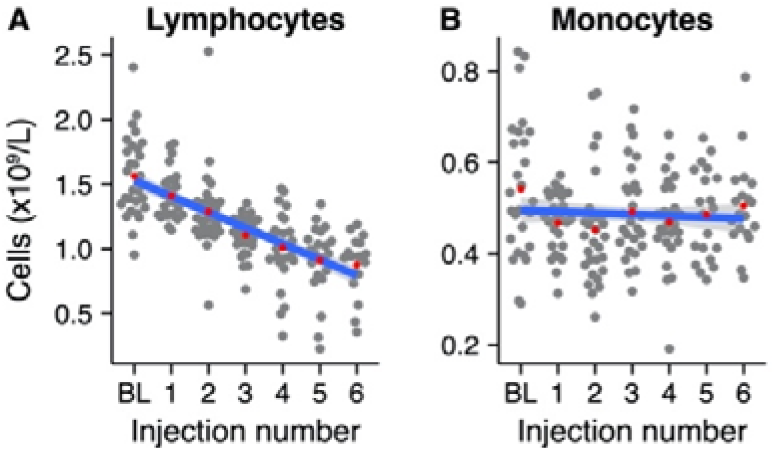
Normalized absolute mononuclear cell counts during Radium-223 treatment. (A) Lymphocyte and (B) monocyte counts at baseline (BL) and after each Radium-223 injection. Red dots indicate the group means. The blue line represents the fitted linear regression model, including 95% CI.

Circulating immune cell populations were analyzed for their abundance in peripheral blood during treatment with radium-223. PBMC-derived percentages of cell populations were pre-processed by a logit-transformation to enable analysis using linear regression models (except for ΔMFI; Figure 2, Supplementary Figures 3 and 6). Before analysis, the percentage/ΔMFI data were normalized per patient, as described above. (Figure 2A). Immune cell subsets were visualized in grouped scatterplots per injection number. A linear model was fitted to each immune cell subset to determine the average change per injection number. Furthermore, checkpoint expression on these cells was also analyzed using the ΔMFI.

**Figure 2:**
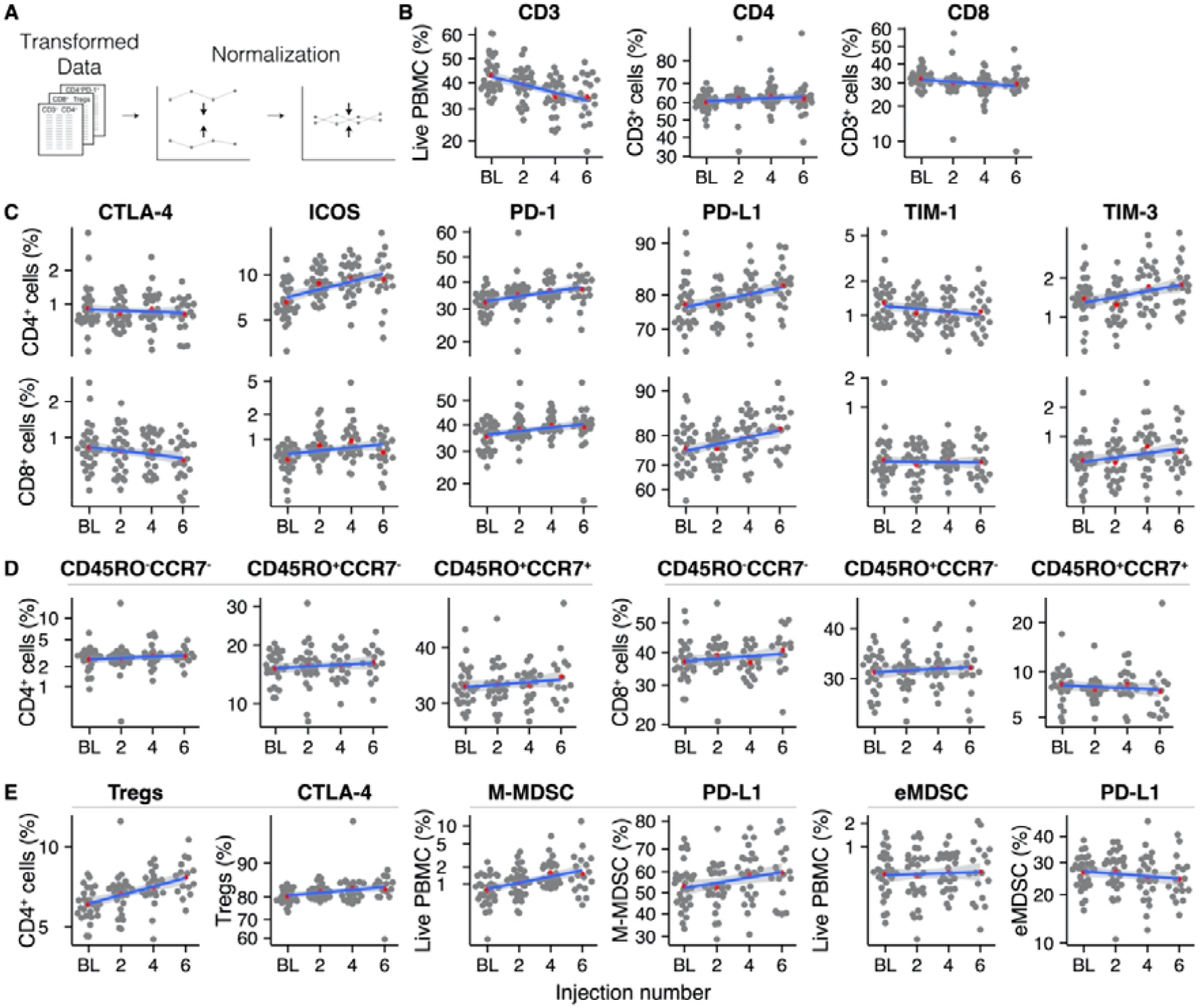
Overview of the analyzed immune cell subsets throughout radium-223 treatment. (A) Data pre-processing steps, (B) CD3^+^, CD4^+^, and CD8^+^ T cells, (C) Immune checkpoint-expressing T cells, (D) Memory and effector T cell subsets, (E) Immunosuppressive cell subsets. Red dots indicate the group means. The blue line represents the fitted linear regression model, including 95% CI.

Next, we quantified longitudinal trends in immune cell subsets as the 6-month change (i.e., change between the subset’s baseline value and the value after the sixth radium-223 injection). We used percentile bootstrapping to determine the 95% confidence interval (CI) of the 6-month change of an immune cell subset over time (unit: percentage change relative to baseline; Figure 3, Supplementary Figures 4 and 7). Specifically, we used 2000 replicates in each bootstrap. Per replicate, we re-sampled the 30 patients with replacement, normalized the data as described above, and fitted a linear model to the sample. Next, the 6-month change between these measures was calculated, including a 95% CI to determine the uncertainty of the bootstrap estimate. Checkpoint expression was analyzed similarly.

**Figure 3:**
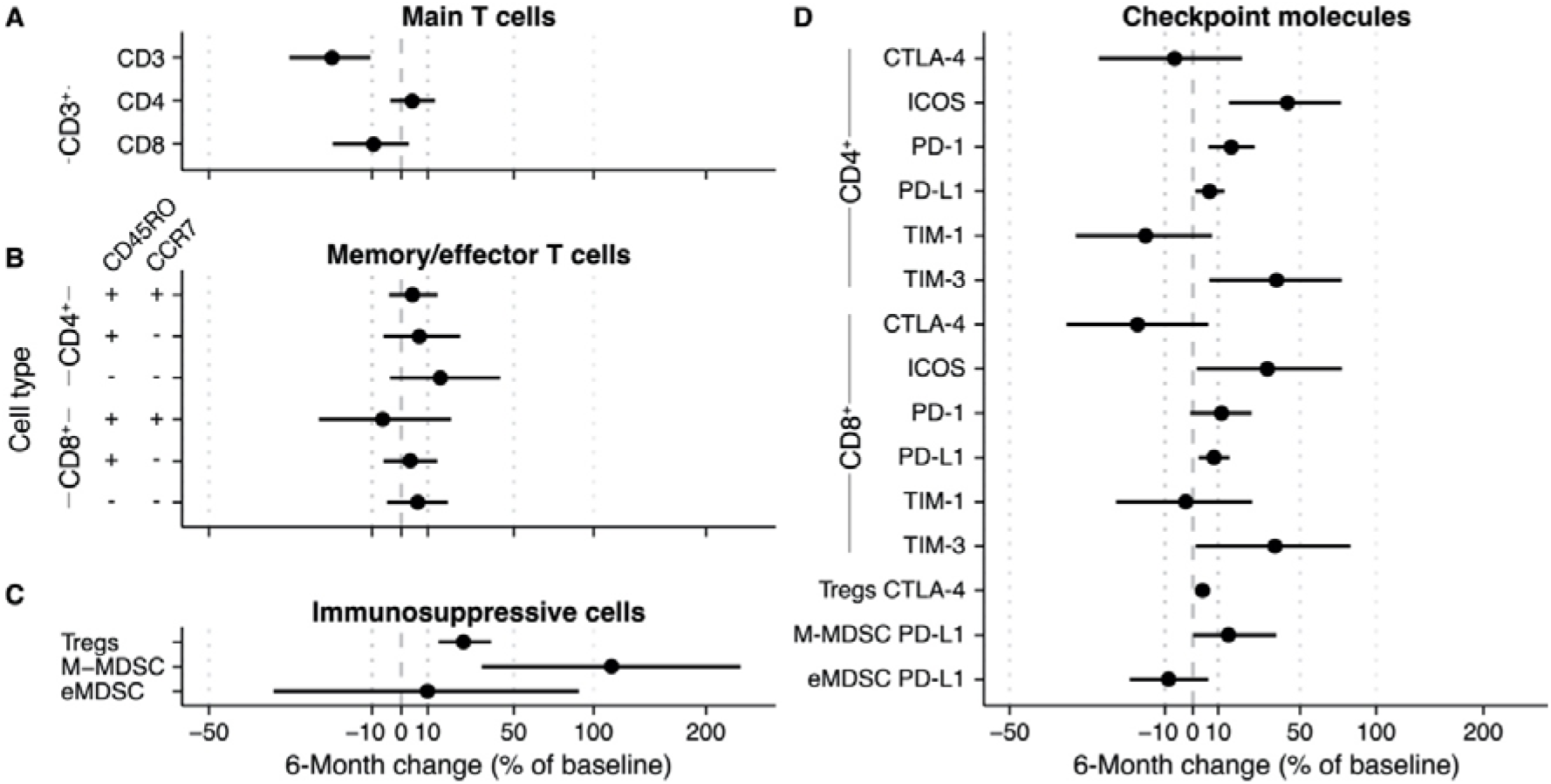
6-Month change estimate of immune cell subsets during radium-223 therapy. The percentage change relative to baseline is calculated using a bootstrap method. Per bootstrap, a linear model is fitted on the logit-transformed and normalized bootstrap sample. Subsequently, the model predictions at baseline and after six months are used to calculate the change in (A) CD3^+^, CD4^+^, and CD8^+^ T cells, (B) memory/effector T cells, (C) immunosuppressive cells, and (D) checkpoint-expressing T cells and monocytes.

In the end, we performed an exploratory subgroup analysis in patients with an ALP response in the same manner.

All R code is available on GitHub: https://github.com/jeroencreemers/Immunophenotyping-radium223-mCRPC.

## RESULTS

### Patient characteristics

A total of 35 mCRPC patients were screened, of which 33 patients met the in- and exclusion criteria (two patients received concomitant enzalutamide). In the final analysis, three patients were excluded because ≤2 injections of radium-223 were administered, and no follow-up blood withdrawal had occurred after discontinuation of treatment. The median age of the study population was 71 years. The median number of prior systemic prostate cancer therapies since diagnosis of metastatic hormone-sensitive prostate cancer was one (range 0-4). Seventeen patients (57%) had received prior docetaxel chemotherapy, and twenty patients (67%) received prior enzalutamide and/or abiraterone. Median baseline ALP was 148 IU/l, and the majority of patients (77%) had high volume (>20) bone metastatic disease. Baseline patient demographics and clinical characteristics are shown in Table 1.

**Table 1.** Baseline patient demographics and clinical characteristics.

|  | All patients (N=30) |  |
| --- | --- | --- |
| Age, years, median (IQR) | 71 | (64-77) |
| Time from mCRPC to radium-223, months, median (IQR) | 23.9 | (10.3-35.3) |
| Gleason score $\geq 8$, n (%) | 21 | (70.0) |
| Extent of disease, n (%) |  |  |
| Low volume ($<6$ bone metastases) | 1 | (3.3) |
| Intermediate volume (6-20 bone metastases) | 4 | (13.3) |
| High volume ($>20$ bone metastases) | 23 | (76.7) |
| Superscan* | 2 | (6.7) |
| Lymph node metastases $\geq 15$ mm | 3 | (10.0) |
| Prior systemic therapies |  |  |
| Median number of prior registered therapies (range) | 1 | (0-4) |
| None, n (%) | 7 | (23.3) |
| Docetaxel, n (%)<sup>†</sup> | 17 | (56.7) |
| Cabazitaxel, n (%) | 3 | (10.0) |
| Abiraterone, n (%) | 12 | (40.0) |
| Enzalutamide, n (%) | 13 | (43.3) |
| Both Abiraterone and Enzalutamide, n (%) | 5 | (16.7) |
| Prior symptomatic skeletal event, n (%) | 13 | (43.3) |
| Opioid use, n (%) | 9 | (30.0) |
| ECOG performance status, n (%) |  |  |
| ECOG 0 | 21 | (70.0) |
| ECOG 1 | 8 | (26.7) |
| ECOG 2 | 1 | (3.3) |
| Hemoglobin, mmol/L, median (IQR) | 7.5 | (7.3-8.0) |
| Platelet count, $\times 10^9$/L, median (IQR) | 239 | (192-275) |
| PSA, ng/ml, median (IQR) | 130 | (53-374) |
| ALP, U/L, median (IQR) | 148 | (100-266) |
| LDH, U/L, median (IQR) | 232 | (203-274) |

ALP = alkaline phosphatase; mCRPC = castration-resistant prostate cancer; ECOG = Eastern Cooperative Oncology Group; IQR = interquartile range; PSA = prostate-specific antigen
\*Superscan refers to a bone scan showing diffuse, intense skeletal uptake of the tracer without renal and background activity.
<sup>†</sup>Including 3 (10%) patients who were treated with upfront docetaxel for metastatic hormone-naïve prostate cancer

Twenty-two (73%) patients received all six radium-223 injections. An ALP decline of ≥30% during therapy was achieved in twenty patients (67%). Four patients (13%) had a PSA decline of ≥30% during therapy. At the time of analysis, 27 patients (90%) were deceased. Median OS was 13.2 months (95% CI 10.2-16.2 months). The majority of patients (60%) showed no new bone metastases on bone scintigraphy after radium-223. Radiological evidence of new pathological lymph node metastases or development of visceral metastases was found in seven patients (26%) and four patients (15%), respectively. Clinical outcomes are shown in Supplementary Table 2.

### Availability of PBMCs for immunophenotyping

PBMCs for immunophenotyping were available of 30, 29, 26, and 20 patients at baseline and after two, four, and six injections of radium-223, respectively (Supplementary Figure S2). Missing data was mostly a consequence of the preterm discontinuation of radium-223 therapy. In one patient (ID-25), no PBMCs were collected after the second injection of radium-223.

### Absolute lymphocyte and monocyte count

We first analyzed the absolute peripheral lymphocyte and monocyte counts to account for potential longitudinal variation in mononuclear cells in the population. We fitted linear regression models to the normalized absolute lymphocyte and monocyte counts per injection number (as a surrogate marker for time). Lymphocyte counts declined by approximately a factor two during treatment with radium-223 (Figure 1A; slope: -0.12 * 10^9^ cells/L per injection number, 95% CI: -0.143 - -0.102), while monocyte counts remained relatively stable over the course of treatment (Figure 1B; slope: -0.003 *10^9^ cells/L per injection number, 95% CI: -0.012 - 0.005).

### Immunophenotype of lymphocyte and monocyte subsets during radium-223 therapy

To investigate longitudinal changes in circulating immune cell subsets in mCRPC patients during treatment with radium-223, we applied two pre-processing steps to the PBMC data: a logit transformation and a normalization step (see Methods; Figure 2A).

We first analyzed the PBMCs for the presence of T cells (CD3^+^ cells; Figure 2B). Despite normalization, substantial variation remained present in the data. Overall, the percentage of T cells within the population of PBMCs decreased during radium-223 treatment. Within the CD3^+^ cells, we distinguished two T cell subsets: CD4^+^ T cells and CD8^+^ T cells (Figure 2B). No major changes were seen in these subsets. Next, we determined the fraction of CD4^+^ and CD8^+^ T cells that expressed costimulatory (OX-40, ICOS, TIM-1) and inhibitory (CTLA-4, LAG-3, PD-L1, PD-1, TIM-3) checkpoint molecules, respectively (Figure 2C). The expression of OX-40 and LAG-3 on both subsets was practically absent; hence these checkpoint molecules were omitted from further analyses. Expression of CTLA-4, TIM-1, and TIM-3 was observed on a small subset (<5%) of CD4^+^ and CD8^+^ T cells, while expression of PD-1 (>30%) and PD-L1 (>70%) was more pronounced. A larger proportion of CD4^+^ cells than of CD8^+^ cells expressed ICOS (6.6% and <1% at baseline, respectively). The fraction of CD4^+^ and CD8^+^ T cells expressing PD-1 and PD-L1, as well as the fraction of CD4^+^ T cells expressing ICOS, increased slightly during treatment, while other checkpoint-expressing subsets remained stable over time. The relative expression of checkpoint molecules on CD4^+^ and CD8^+^ cells (i.e., ΔMFI) is shown in Supplementary Figure S3.

Based on the expression of memory marker CD45RO and lymphoid tissue homing chemokine receptor CCR7, we identified central memory (CD45RO^+^CCR7^+^), effector memory (CD45RO^+^CCR7^-^), and effector (CD45RO^-^CCR7^-^) cells within the CD4^+^ and CD8^+^ T cell subsets (Figure 2D). All memory and/or effector phenotypes within the CD4^+^ and the CD8^+^ subset appeared stable throughout radium-223 therapy.

Subsequently, we studied four immunosuppressive cell types: regulatory T cells (Tregs; CD3^+^CD4^+^CD25^+^FoxP3^+^), monocytic MDSCs (M-MDSCs; CD11b^+^CD14^+^CD15^-^HLA-DR^low/-^), polymorphonuclear MDSCs (PMN-MDSCs; CD11b^+^CD14^-^CD15^+^HLA-DR^low/-^), and early MDSCs (eMDSC; lin^-^CD14^-^CD15^-^CD33^+^HLA-DR^low/-^). PMN-MDSCs were not detected, likely because we used cryopreserved samples. While the portion of M-MDSCs and eMDSCs, as well as the subsets expressing PD-L1, remained stable during treatment, we observed a small increase in the Treg fraction (Figure 2E).

### The uncertainty associated with the longitudinal changes in immune cell subsets

To measure the degree of sampling variation in the described longitudinal changes of immune cell subsets during radium-223 treatment, we used a bootstrapping approach.

We applied bootstrapping to the main T cell subsets (Figure 3A), the memory/effector T cell subsets (Figure 3B), and the immunosuppressive subsets (Figure 3C). With regard to the main T cell subsets, the bootstrap approach estimated an overall 6-month decrease of 20.3% (CI -31.8% - -8.8%) of the CD3^+^ subset (Figure 3A).

The changes in the CD4^+^ and CD8^+^ fractions of the CD3^+^ subset were uncertain (Figure 3A). In the memory/effector T cell phenotypes, the largest change estimate was observed in the CD4^+^CD45RO^-^ CCR7^-^ subset (bootstrap estimate +15.1%, CI -4% – 42.9%). All other memory/effector phenotypes had less pronounced change estimates with CIs spanning across both positive and negative changes (Figure 3B). As indicated in Figure 3C, the proportion of Tregs in peripheral blood seemed to increase over time (25.1%; CI 14.3% - 38.2%). While the bootstrap estimate indicated an increase in the fraction of M-MDSCs during therapy, the CI was wide (113.3%; CI 33.6% - 239.6%). In contrast to the other two immunosuppressive subsets, the eMDSC change estimate was inconclusive (9.9%; -36.9% - 89.7%).

Concerning the checkpoint-expressing subsets, we observed a pronounced increase in the ICOS- and TIM-3-expressing proportion of CD4^+^ and CD8^+^ T cells, albeit with wide CIs (Figure 3D; numerical values are added as Supplementary Table S3). Less pronounced increases were seen in the fractions of PD-1- and PD-L1-expressing CD4^+^ and CD8^+^ T cells and PD-L1-expressing M-MDSCs. For the CTLA-4- and TIM-1-expressing subsets, changes were small (i.e., CD4^+^CTLA-4^+^ and CD8^+^TIM-1^+^), and/or the sampling variation was large (Figure 3D). Interestingly, while the fraction of Tregs increased over time, the proportion of CTLA-4-expressing Tregs hardly changed.

In addition, the 6-month change in the relative expression of checkpoint molecules on lymphocytes and monocytes, as analyzed with the ΔMFI, followed the same trend as the immune cell counts. The 6-month change estimates of the ΔMFI ranged from 0-10% (Supplementary Figure S4; Supplementary Table 4).

### Subgroup analysis in patients with an ALP response

Since our data indicated longitudinal changes in fractions of peripheral lymphocyte or monocyte subsets in the entire study population, and ALP changes are considered a surrogate marker for response to radium-223, we hypothesized that potential changes might be more pronounced in patients with an ALP response to radium-223 therapy (n=20; 67%). PBMCs for immunophenotyping were available on all time points for thirteen patients in this subset (Supplementary Figure S5). Visually, there is a strong resemblance between the subgroup with ALP responders and the entire population in terms of the occurrence of cells, their distribution, and time trends (Supplementary Figure S6). The bootstrapping approach supports this resemblance: we observed similar trends compared to the entire population, with higher uncertainty due to the smaller sample size (Supplementary Figure S7; Supplementary Table S5). Therefore, our data do not support a more pronounced induction of fractions of immune cell subsets over time in patients with an ALP response, compared to those without a biochemical response to radium-223 therapy.

## DISCUSSION

In this prospective exploratory study, we performed a comprehensive evaluation of the immunological changes during radium-223 therapy by phenotyping PBMCs of 30 mCRPC patients. Overall, a substantial decrease in absolute lymphocyte counts was observed. While the total lymphocyte count decreased, we observed an increase in the proportion of T cells that expressed costimulatory (ICOS) or inhibitory (PD-L1, PD-1 and TIM-3) checkpoint molecules. In the immunosuppressive subsets, the proportion of Tregs and M-MDSCs increased during this study. A subgroup analysis in ALP responders indicated that the observed changes were not more pronounced in responding patients.

We observed a nearly two-fold decrease in absolute lymphocyte counts. In line with this, the fraction of CD3^+^ T cells in PBMCs decreased. Although radium-223 is known to induce hematologic toxicity (neutropenia, thrombopenia) in a subset of patients, lymphopenia is not considered a common side effect of radium-223. Lymphopenia, defined as lymphocyte counts ≤ 0.8*10^9^, was reported in only 1% of patients in the ALSYMPCA trial [16], whereas seventeen patients (56.7%) in our study developed lymphopenia during treatment. A possible explanation for the higher incidence of lymphopenia is the higher prevalence of patients with extensive bone metastases (i.e., >20 bone metastases in 83% of patients). In the pivotal ALSYMPCA trial, only 41% of patients had high volume bone metastatic disease. The decrease in lymphocyte counts might be a consequence of the direct cytotoxic effects of radium-223 on the bone marrow in patients with an already impaired bone marrow due to extensive bone metastases [17]. It is important to acknowledge that we only investigated circulating immune cells. We did not investigate changes in tumor-infiltrating immune cells. Therefore, it is unclear how the decrease in peripheral lymphocyte counts affects the number of tumor-infiltrating lymphocytes.

An increase was observed in the proportion of CD4^+^ and CD8^+^ T cells expressing immune checkpoint molecules PD-L1, ICOS, PD-1, or TIM-3. Checkpoint molecules play an essential role in the regulation of immune cell activity. ICOS is a costimulatory checkpoint molecule that enhances T cell activation via binding to its ligand on antigen-presenting cells. PD-L1, PD-1, and TIM-3 are inhibitory checkpoint molecules. These checkpoint molecules inhibit T cell proliferation and activation to limit the immune response and maintain immune homeostasis. Although inhibitory checkpoint molecules are often considered markers of immune exhaustion, inhibitory checkpoint molecules are upregulated upon immune cell activation and are, therefore, also markers of immune activation [18,19]. Only one study in fifteen mCRPC patients has reported changes in checkpoint molecule expression during radium-223. Herein, PD-1^+^ effector memory CD8^+^ T cells decreased [8], while we observed a small increase in total CD8^+^PD-1^+^ T cells. In line with our findings, studies with ionizing radiation show upregulation of checkpoint molecule expression in the tumor microenvironment. Several mice studies have described that PD-1 [20] and PD-L1 [21–23] are upregulated in the tumor microenvironment following irradiation and that combining irradiation with anti-PD-(L)1 improved tumor control compared to either therapy alone. Oweida and colleagues observed an increase in ICOS and TIM-3 expression on tumor-infiltrating T cells during treatment with radiotherapy and PD-L1 blockade [24]. Moreover, a recent study in patients with head and neck cancer showed that PD-1 and CTLA-4 expression on PBMCs increased following radiation therapy [9].

Besides the increase in the fraction of checkpoint molecule-expressing T cells, we observed an increase in the proportion of two immunosuppressive subsets during radium-223 therapy (i.e., Tregs and M-MDSCs). There is no data on the effect of radium-223 on Tregs or M-MDSCs, but several studies have reported that ionizing radiation can lead to the accumulation of circulating and tumor-infiltrating Tregs [9,24–26] and MDSCs [27], supporting our findings here.

It is unclear how the changes observed during radium-223 affect antitumor immunity. We hypothesized that radium-223 would lead to immune cell activation. However, except for the increase in the fraction of ICOS-expressing T cells, our findings – specifically, the relative increase in Tregs and M-MDSCs and the upregulation of inhibitory checkpoints molecules – are associated with immune suppression. It is possible that the relative increase in Tregs, M-MDSCs, and checkpoint-expressing T cells prevents excessive immune activity during radium-223 therapy or reflects the migration of (non-exhausted) effector T cells into the tumor [28]. Given these hypotheses, it might be effective to combine radium-223 with immunotherapy due to synergistic effects on the immune system. Another possibility is that the relative increase in immunosuppressive cells and the upregulation of inhibitory checkpoint molecules during radium-223 therapy abrogate the immune- promoting effects of radium-223 and inhibit an effective antitumor immune response. In the latter case, treatment strategies combining radium-223 with immune-based therapies might not be effective unless we are informed about the most critical immunosuppressive mechanisms in these patients and can overcome them.

Knowledge of the immunological effects of radium-223 is vital to improve the care for patients with mCRPC. Although Sipuleucel-T – a cellular immunotherapy – is registered for the treatment of mCRPC [29], checkpoint inhibitor monotherapy could not induce clinically meaningful responses in unselected cohorts of mCRPC patients [30–32]. While checkpoint inhibitor monotherapy is not the way forward for all mCRPC patients, checkpoint inhibitors may be of value in specific subgroups (NCT04104893) [33–35] or in combination with other therapies (NCT02861573). Subgroups of interest include patients with a high tumor mutational burden [34] or DNA damage repair deficiency [35–37]. The data on combination strategies of radium-223 with immunotherapy is scarce. In a randomized phase II trial, including 32 mCRPC patients, the combination of sipuleucel-T with radium-223 was found to increase median progression-free survival compared to sipuleucel-T alone (10.7 versus 3.1 months; HR 0.35, 95% CI 0.15-0.81; p=0.02). PSA responses were more frequently observed in the combination arm (33% versus 0%) [38], supporting the idea that radium-223 promotes antitumor immunity. Results from a recent single-arm, phase Ib trial indicated a limited efficacy of radium-223 in combination with PD-L1 inhibitor atezolizumab, with confirmed objective response rates of only 6.8% and PSA responses in 4.5% of patients [39]. Other phase II trials combining radium-223 with checkpoint inhibitors are ongoing (NCT02814669, NCT03093428, NCT04109729, NCT04071236).

This study has some limitations. We did not include a control arm consisting of mCRPC patients that did not receive radium-223 therapy. Therefore, it remains uncertain if the observed immunological changes are indeed a result of radium-223 therapy. The observed changes may be a consequence of disease progression rather than an effect of radium-223. Another constraint is that we did not investigate changes in tumor-infiltrating immune cells. Therefore, it remains unclear whether changes in the composition and abundance of circulating immune cells reflect changes in tumor-infiltrating lymphocytes. It is difficult to interpret our results without knowledge of changes in tumor-infiltrating immune cells. Further research should uncover how the observed changes in PBMCs correlate with changes in the tumor microenvironment. A third limitation is the small sample size relative to the number of markers being studied.

In summary, we observed a decrease in absolute lymphocyte counts and an increase in the proportion of checkpoint-expressing T cells, Tregs, and M-MDSCs. Our findings provide initial insights into the temporal dynamics of immune cell subsets in mCRPC patients and guide further research into radium-223-induced immune mechanisms. A thorough understanding of these mechanisms might pave the way toward optimizing treatment timing and effective combination strategies.

## Data Availability

All data and code is available on GitHub.

https://github.com/jeroencreemers/Immunophenotyping-radium223-mCRPC

## ACKNOWLEDGEMENTS

We thank all patients and their families who participated in this study. We thank Merijn Janssen, Marga Ouwens, Marjo van de Ven, Maarten de Bakker, and Maarten Vinken for the collection of the blood samples.

