## Supplementary Material for "Immunophenotyping reveals longitudinal changes in circulating immune cells during radium-223 therapy in patients with metastatic castration-resistant prostate cancer"

**SUPPLEMENTARY TABLES**

**Supplementary Table 1 – Antibody details**

| **Panel 1 (T cell panel)** | | | | | |
| --- | --- | --- | --- | --- | --- |
| **Antigen** | **Clone** | **Conjugate** | **Company** | **Catalog No.** | **Dilution** |
| CD3 | UCHT1 | BV605 | Biolegend | 300460 | 1:40 |
| CD4 | SK3 | BV510 | BD biosciences | 659454 | 1:25 |
| CD8 | SK1 | PerCP-Cy5.5 | BD biosciences | 565310 | 1:10 |
| CD25 | M-A251 | BV421 | BD biosciences | 562443 | 1:40 |
| CD45RO | UCHL1 | PE-Cy7 | BD biosciences | 560608 | 1:25 |
| CTLA-4 | BNI3 | PE | BD biosciences | 557301 | 1:10 |
| CCR7 | 150503 | AF647 | BD biosciences | 560816 | 1:5 |
| FoxP3 | PCH101 | A488 | eBioscience | **53-4776-42** | 1:10 |
| **Panel 2 (Checkpoint panel 1)** | | | | | |
| **Antigen** | **Clone** | **Conjugate** | **Company** | **Catalog No.** | **Dilution** |
| CD3 | UCHT1 | PE-Cy7 | eBioscience | 25-0038-42 | 1:25 |
| CD4 | SK3 | BV510 | BD Biosciences | 659454 | 1:25 |
| CD8 | G42-8 | BV605 | BD Biosciences | 743066 | 1:40 |
| LAG-3 | 3DS223H | FITC | eBioscience | 11-2239-42 | 1:10 |
| PD-L1 | 29E.2A3 | APC | BioLegend | 329708 | 1:5 |
| ICOS | DX29 | PerCP-Cy5.5 | BD Bioscience | 562833 | 1:10 |
| PD-1 | EH12.2H7 | BV421 | BioLegend | 329920 | 1:5 |
| TIM-3 | F38-2E2 | PE | BioLegend | 345006 | 1:10 |
| **Panel 3 (Checkpoint panel 2)** | | | | | |
| **Antigen** | **Clone** | **Conjugate** | **Company** | **Catalog No.** | **Dilution** |
| CD3 | UCHT1 | BV605 | Biolegend | 300460 | 1:40 |
| CD4 | SK3 | BV510 | BD Biosciences | 659454 | 1:25 |
| CD8 | SK1 | PerCP-Cy5.5 | BD Biosciences | 565310 | 1:10 |
| OX-40 | ACT35 | FITC | BD biosciences | 555837 | 1:5 |
| CTLA-4 | BNI3 | PE | Biosciences | 557301 | 1:10 |
| PD-L1 | MIH1 | PE-Cy7 | Biosciences | 558017 | 1:5 |
| PD-1 | EH12.2H7 | BV421 | BioLegend | 329708 | 1:5 |
| TIM-1 | 1D12 | APC | Biolegend | 353906 | 1:25 |
| **MDSC panel** | | | | | |
| **Antigen** | **Clone** | **Conjugate** | **Company** | **Catalog No.** | **Dilution** |
| CD3 | HIT3α | FITC | BD Biosciences | 555339 | 1:10 |
| CD11b | ICRF44 | BV605 | BD Biosciences | 562723 | 1:10 |
| CD14 | MΦP9 | BV421 | BD Biosciences | 560827 | 1:10 |
| CD15 | HI98 | PerCP-Cy5.5 | BD Biosciences | 560827 | 1:10 |
| CD19 | HD37 | FITC | Agilent | F0768 | 1:10 |
| CD20 | L27 | FITC | BD Biosciences | 345792 | 1:10 |
| CD33 | WM53 | APC | Biolegend | 303408 | 1:10 |
| CD56 | B159 | FITC | BD Biosciences | 562794 | 1:10 |
| PD-L2 | MIH18 | PE | BD Biosciences | 558066 | 1:5 |
| PD-L1 | MIH1 | PE-Cy7 | BD Biosciences | 558017 | 1:5 |
| HLA-DR | G46-6 | BV510 | BD Biosciences | 563083 | 1:25 |

**Supplementary Table 2 – Clinical outcomes of radium-223 therapy**

|  | | All patients (N=30) | |
| --- | --- | --- | --- |
| Number of received injections, n (%) | |  |  |
|  | 3 injections | 2 | (6.7) |
|  | 4 injections | 3 | (10.0) |
|  | 5 injections | 3 | (10.0) |
|  | 6 injections | 22 | (73.3) |
| Biochemical response evaluation | |  |  |
|  | PSA decline ≥30%, n (%) | 4 | (13.3) |
|  | ALP decline ≥30%, n (%) | 20 | (66.7) |
|  | ALP normalization*, n (%) | 10 | (50.0) |
| Radiological evaluation on bone scintigraphy^†^ | |  |  |
|  | Stable number of bone metastases, n (%) | 12 | (60.0) |
|  | New bone metastases, n (%) | 8 | (40.0) |
| Radiological evaluation on CT^‡^ | | | |
|  | New lymph node metastases, n (%) | 7 | (25.9) |
|  | New visceral metastases, n (%) | 4 | (14.8) |
| Overall survival, months, median (95% CI) | | 13.2 | (10.2-16.2) |
| *ALP = alkaline phosphatase; CI = confidence interval; CT = computer tomography; PSA = prostate-specific antigen* | | | |
| ** If elevated alkaline phosphatase (e.g. ≥ 115 U/l) at baseline*  *^†^ 20 (67%) patients underwent bone scintigraphy after radium-223*  *^‡^ 27 (90%) patients underwent CT of thorax, abdomen and pelvis after radium-223* | | | |

**Supplementary Table 3 – Bootstrap estimates and confidence interval of all cell subsets**

|  | Cell subset | 6-month change  (% of baseline) | Lower bound of CI | Upper bound of CI |
| --- | --- | --- | --- | --- |
|  | CD3 | -22.1 | -33.2 | -10.5 |
|  | CD14 | 27.2 | 13.2 | 42.8 |
| CD4^+^ | Total | 4 | -3.9 | 12.9 |
|  | CD45RO^-^CCR7^-^ | 15.1 | -4 | 42.9 |
|  | CD45RO^+^CCR7^-^ | 6.7 | -6.2 | 23.7 |
|  | CD45RO^+^CCR7^+^ | 4.1 | -4.3 | 14 |
|  | CTLA-4 | -6.7 | -30 | 20.4 |
|  | PD-L1 | 6.5 | 0.9 | 12.7 |
|  | ICOS | 43 | 14.6 | 75.1 |
|  | PD-1 | 15.6 | 6 | 26.3 |
|  | TIM-1 | -16.4 | -35.8 | 7.5 |
|  | TIM-3 | 37.2 | 6.4 | 75.6 |
| CD8^+^ | Total | -9.5 | -21.9 | 2.7 |
|  | CD45RO^-^CCR7^-^ | 6.1 | -5 | 18.3 |
|  | CD45RO^+^CCR7^-^ | 3.3 | -6.2 | 13.9 |
|  | CD45RO^+^CCR7^+^ | -6.5 | -25.7 | 19.7 |
|  | CTLA-4 | -18.9 | -38 | 6 |
|  | PD-L1 | 8.3 | 2.2 | 14.9 |
|  | ICOS | 32.5 | 1.5 | 75.7 |
|  | PD-1 | 11.4 | -1 | 24.8 |
|  | TIM-1 | -2.7 | -25.2 | 25.1 |
|  | TIM-3 | 36.4 | 1.1 | 81.5 |
|  | Tregs | 25.1 | 14.3 | 38.2 |
|  | Tregs CTLA | 3.7 | 1.3 | 6.1 |
|  | M-MDSC | 113.3 | 33.6 | 239.6 |
|  | M-MDSC PD-L1 | 14.4 | 0.1 | 37.1 |
|  | eMDSC | 9.9 | -36.9 | 89.7 |
|  | eMDSC PD-L1 | -8.8 | -21.3 | 6 |

**Supplementary Table 4 – Bootstrap estimates and confidence interval of the ∆MFI**

|  | Cell subset | 6-month change  (% of baseline) | Lower bound of CI | Upper bound of CI |
| --- | --- | --- | --- | --- |
| CD4^+^ | CTLA-4 | 0.3 | -0.3 | 0.9 |
|  | PD-L1 | 7.1 | 2.2 | 11.7 |
|  | ICOS | 4.5 | 1.5 | 7.3 |
|  | PD-1 | 10.1 | -4.7 | 23.2 |
|  | TIM-1 | -0.1 | -0.3 | 0.2 |
|  | TIM-3 | 4.9 | 2.5 | 7.3 |
| CD8^+^ | CTLA-4 | -0.3 | -0.8 | 0.3 |
|  | PD-L1 | 7.9 | 2.7 | 12.9 |
|  | ICOS | 7.1 | 2.5 | 12.9 |
|  | PD-1 | 7.3 | -15.7 | 32.5 |
|  | TIM-1 | -0.3 | -1.4 | 0.7 |
|  | TIM-3 | 5.6 | 1.9 | 9.1 |
|  | Tregs CTLA-4 | 4.9 | 0.1 | 9.7 |
|  | M-MDSC PD-L1 | 6.3 | -1.3 | 14.4 |
|  | eMDSC PD-L1 | -0.1 | -3.8 | 3.9 |

**Supplementary Table 5 – Bootstrap estimates and confidence interval of the ALP-responding subgroup**

|  | Cell subset | 6-month change  (% of baseline) | Lower bound of CI | Upper bound of CI |
| --- | --- | --- | --- | --- |
|  | CD3 | -17.1 | -30.1 | -3.7 |
|  | CD14 | 16.4 | 2.1 | 31.4 |
| CD4^+^ | Total | 1.7 | -9.6 | 17.4 |
|  | CD45RO^-^CCR7^-^ | 19 | -3.7 | 46.3 |
|  | CD45RO^+^CCR7^-^ | 8.5 | -9 | 29.2 |
|  | CD45RO^+^CCR7^+^ | 2.4 | -8.7 | 15.7 |
|  | CTLA-4 | -16.1 | -43.5 | 17.3 |
|  | PD-L1 | 4.2 | -3.1 | 11.4 |
|  | ICOS | 26 | -5 | 67.6 |
|  | PD-1 | 12.9 | -0.7 | 26.2 |
|  | TIM-1 | -21.4 | -46.8 | 6 |
|  | TIM-3 | 30.9 | -7.8 | 85.7 |
| CD8^+^ | Total | -6.8 | -23.4 | 9.3 |
|  | CD45RO^-^CCR7^-^ | 4.5 | -9.9 | 22.1 |
|  | CD45RO^+^CCR7^-^ | 8.8 | -2.4 | 20.7 |
|  | CD45RO^+^CCR7^+^ | -8.8 | -34.5 | 27.9 |
|  | CTLA-4 | -23.1 | -45.1 | 8.1 |
|  | PD-L1 | 6.7 | -1.3 | 15.7 |
|  | ICOS | 8 | -17 | 39.7 |
|  | PD-1 | 6.9 | -7.9 | 22.2 |
|  | TIM-1 | -8.5 | -35.7 | 28.3 |
|  | TIM-3 | 13.3 | -18.6 | 49.9 |
|  | Tregs | 25.1 | 11.6 | 42 |
|  | Tregs CTLA | 2.6 | 0 | 5.6 |
|  | M-MDSC | 111.9 | 23.3 | 286 |
|  | M-MDSC PD-L1 | 16.4 | -2.8 | 52.6 |
|  | eMDSC | 14.2 | -44.3 | 142.6 |
|  | eMDSC PD-L1 | -8.9 | -23.5 | 10.3 |

**SUPPLEMENTARY FIGURES**

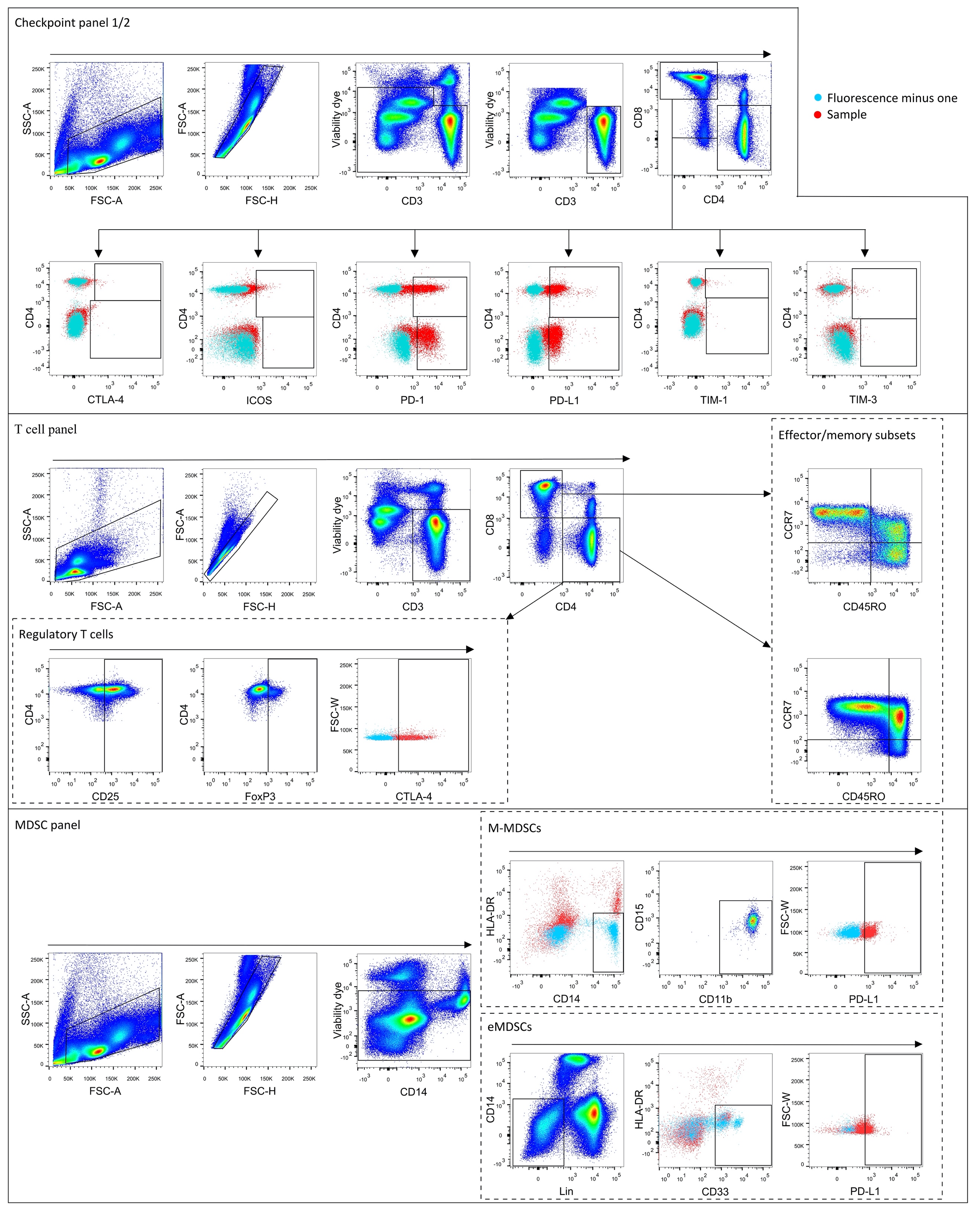

**Supplementary figure 1: Gating strategy.**

(A) Checkpoint panel 1 and 2. Gates for checkpoint expression on CD4^+^ (upper gate) and CD8^+^ cells (lower gate) are shown in the same graph. (B) T cell panel (C) MDSC panel. Fluorescence minus one (FMO) controls were used to gate population that expressed checkpoint molecules and HLA-DR.

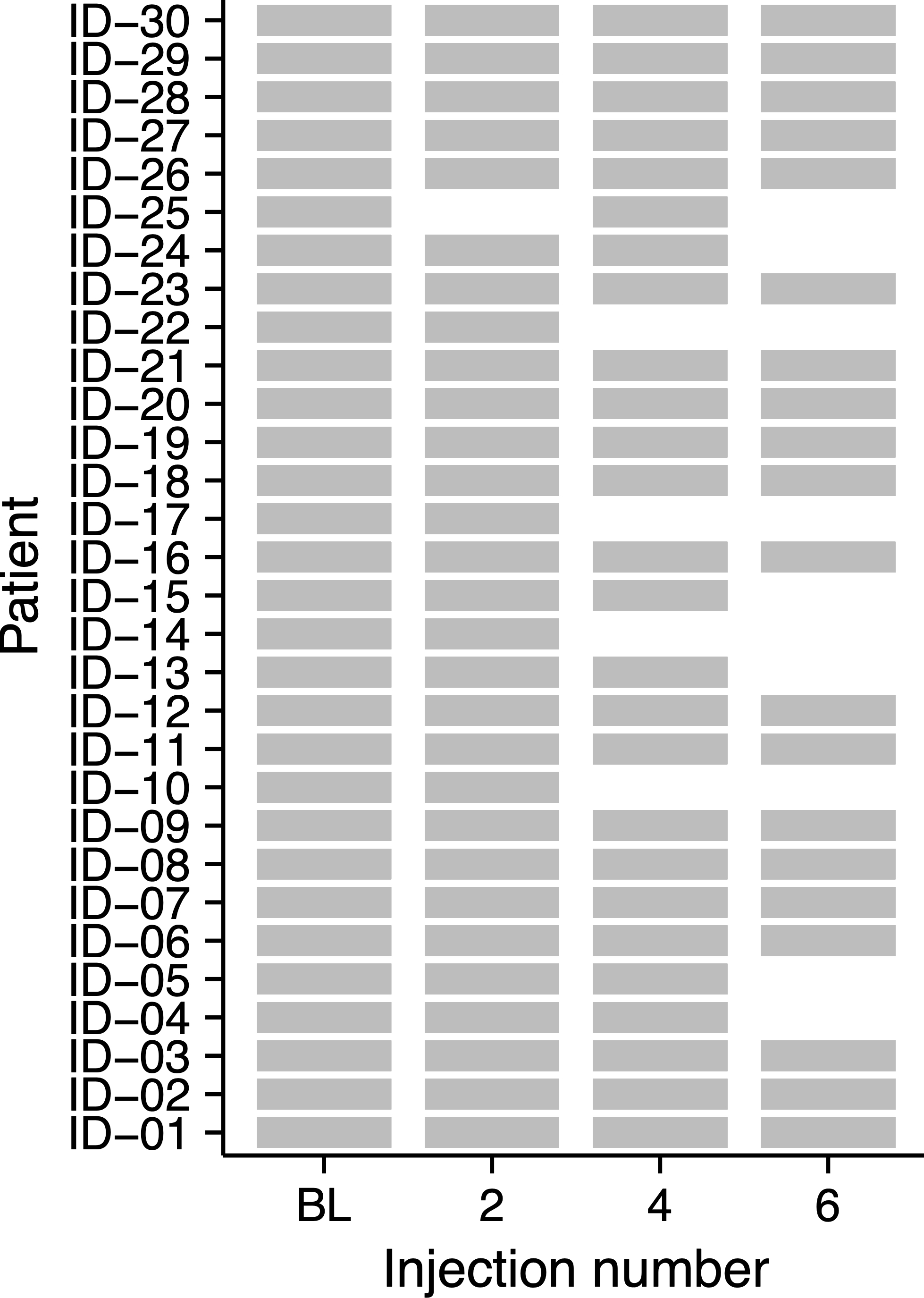

**Supplementary figure 2: Availability of PBMCs for immunophenotyping per injection.**

A grey rectangle indicates the availability of PBMCs at baseline (BL), or after the 2^nd^, 4^th^, or 6^th^ Radium-223 injection.

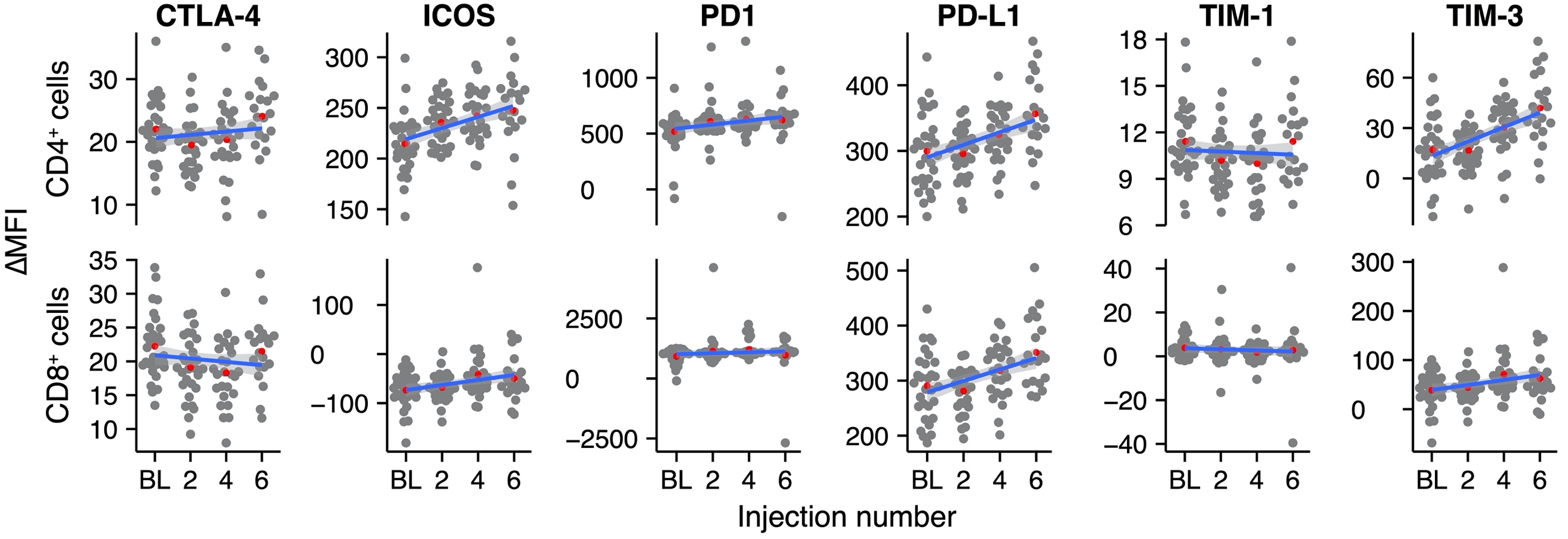

**Supplementary figure 3: Analysis of the ∆MFI of checkpoint molecules on CD4^+^ and CD8^+^ T cells.**

∆MFI values are normalized. Red dots indicate the group mean. The blue line represents the fitted linear regression model, including 95% CI.

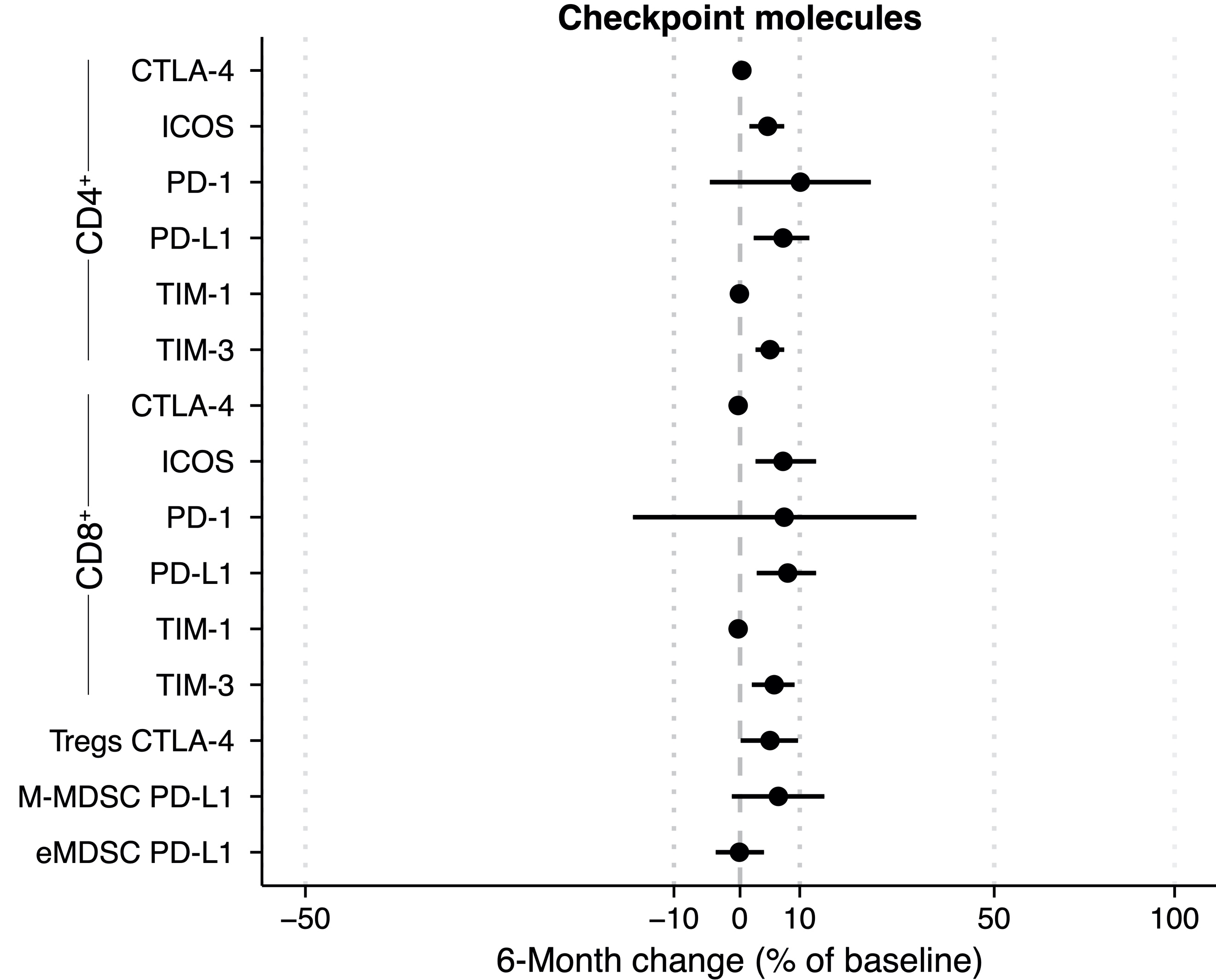

**Supplementary figure 4: 6-Month change estimates of the ∆MFI of the checkpoint molecules (bootstrap approach).**
The percentage change relative to baseline is calculated using a bootstrap method. Per bootstrap, a linear model is fitted on the logit-transformed and normalized bootstrap sample. Subsequently, the model predictions at baseline and after six months are used to calculate the change in checkpoint-expressing T cells over time.

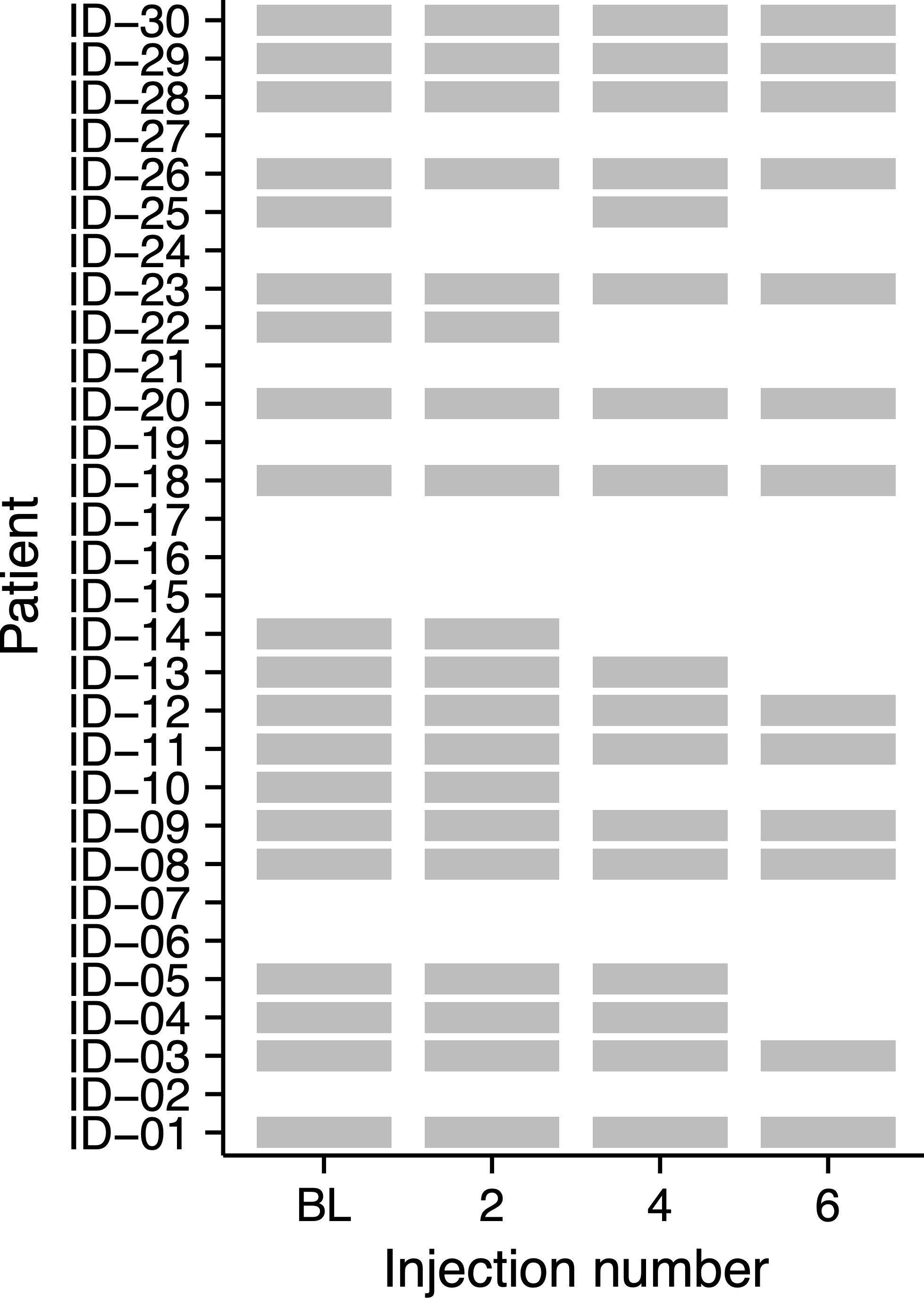

**Supplementary figure 5: Availability of PBMCs for immunophenotyping in the ALP responding subgroup per injection.**

A grey rectangle indicates the availability of PBMCs at baseline (BL), or after the 2^nd^, 4^th^, or 6^th^ Radium-223 injection.

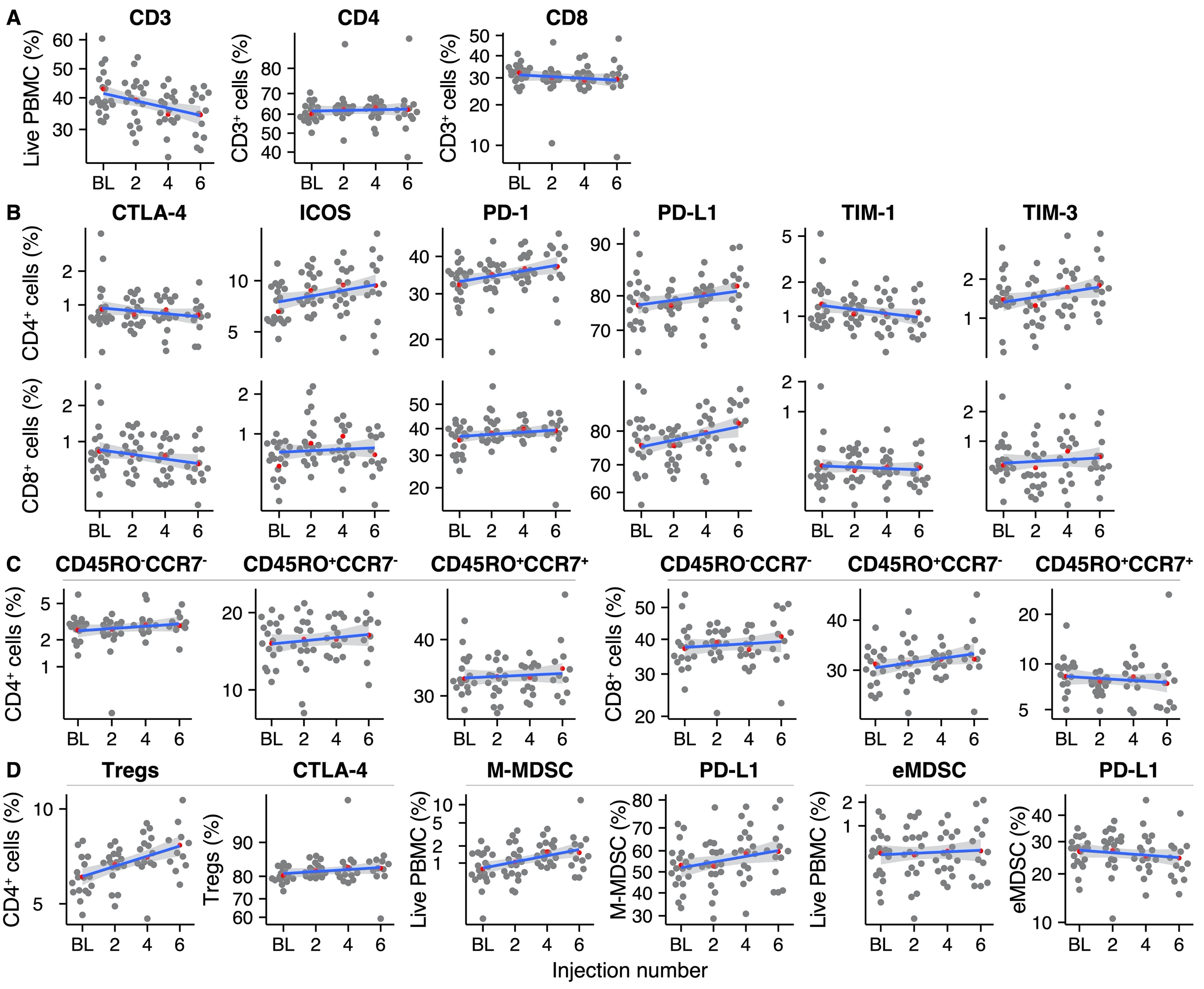

**Supplementary figure 6: Overview of the immune cell subsets in the ALP responding subgroup throughout radium-223 treatment.** (A) CD3^+^, CD4^+^, and CD8^+^ T cells, (B) Immune checkpoint-expressing T cells, (C) Memory and effector T cell subsets, (D) Immunosuppressive cell subsets. Red dots indicate the group means. The blue line represents the fitted linear regression model, including 95% CI.

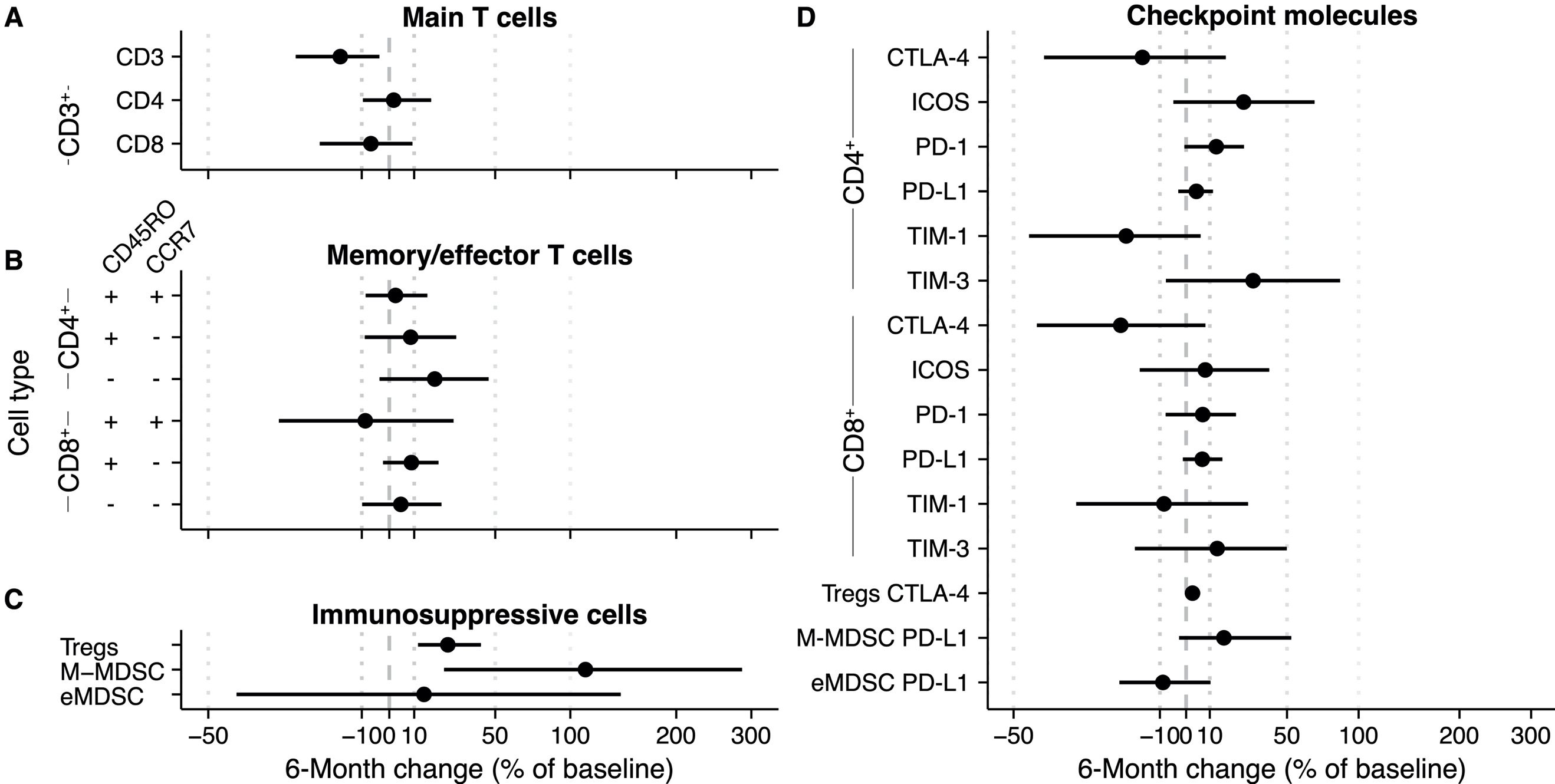

**Supplementary figure 7: Subgroup analysis: 6-Month change estimate of immune cell subsets during radium-223 therapy in ALP responding patients.** The percentage change relative to baseline is calculated using a bootstrap method. Per bootstrap, a linear model is fitted on the logit-transformed and normalized bootstrap sample. Subsequently, the model predictions at baseline and after six months are used to calculate the change in (A) CD3^+^, CD4^+^, and CD8^+^ T cells, (B) memory/effector T cells, (C) immunosuppressive cells, and (D) checkpoint-expressing T cells and monocytes.
